# Tocilizumab in nonventilated patients hospitalized with Covid-19 pneumonia

**DOI:** 10.1101/2020.10.21.20210203

**Authors:** Carlos Salama, Jian Han, Linda Yau, William G. Reiss, Benjamin Kramer, Jeffrey D. Neidhart, Gerard J. Criner, Emma Kaplan-Lewis, Rachel Baden, Lavannya Pandit, Miriam L. Cameron, Julia Garcia-Diaz, Victoria Chávez, Martha Mekebeb-Reuter, Ferdinando Lima Menezes, Reena Shah, Maria F. González-Lara, Beverly Assman, Jamie Freedman, Shalini V. Mohan

## Abstract

**Background:** Coronavirus disease 2019 (Covid-19) pneumonia is often associated with hyperinflammation. Safety and efficacy of the anti–interleukin-6 receptor antibody tocilizumab was evaluated in patients hospitalized with Covid-19 pneumonia.

**Methods:** Nonventilated patients hospitalized with Covid-19 pneumonia were randomized (2:1) to tocilizumab (8 mg/kg intravenous) or placebo plus standard care. Sites enrolling high-risk and minority populations were emphasized. The primary endpoint was cumulative proportion of patients requiring mechanical ventilation or who had died by Day 28.

**Results:** Of 389 randomized patients, 249 patients received tocilizumab and 128 received placebo in the modified intent-to-treat population (Hispanic/Latino, 56.0%; Black/African American, 14.9%; American Indian/Alaska Native, 12.7%; White, 12.7%; other/unknown, 3.7%). The cumulative proportion (95% confidence interval [CI]) of patients requiring mechanical ventilation or who had died by Day 28 was 12.0% (8.52% to 16.86%) and 19.3 % (13.34% to 27.36%) for the tocilizumab and placebo arms, respectively (log-rank P=0.0360; hazard ratio, 0.56 [95% CI, 0.33 to 0.97]). Median time to clinical failure up to Day 28 favored tocilizumab over placebo (hazard ratio 0.55 [95% CI, 0.33 to 0.93]). All-cause mortality by Day 28 was 10.4% with tocilizumab and 8.6% with placebo (weighted difference, 2.0% [95% CI, – 5.2% to 7.8%). In the safety population, serious adverse events occurred in 15.2% of tocilizumab patients (38/250 patients) and 19.7% of placebo patients (25/127).

**Conclusions:** This trial demonstrated the efficacy and safety of tocilizumab over placebo in reducing the likelihood of progression to requiring mechanical ventilation or death in nonventilated patients hospitalized with Covid-19 pneumonia.

**Trial registration:** ClinicalTrials.gov NCT04372186

## INTRODUCTION

Coronavirus disease 2019 (Covid-19) emerged in China in December 2019 and rapidly became a public health emergency.^1,2^ In severe and critical cases, which occur in 14% and 5% of patients, respectively, Covid-19–associated pneumonia can lead to acute respiratory distress syndrome^3,4^; respiratory failure is among the leading causes of death in patients with Covid-19.^5,6^

Patients hospitalized with Covid-19 pneumonia often require invasive mechanical ventilation, and the mortality rate is higher in these patients, especially in those who are > 65 years.^7-10^ In the absence of approved therapies for Covid-19, the mainstay of treatment for patients with Covid-19 pneumonia is symptomatic and supportive,^11^ with dexamethasone being the only therapy demonstrated to improve patient outcomes thus far.^12^ A clear unmet need exists for therapies to treat Covid-19 pneumonia.^3,7^

Therapies for Covid-19 pneumonia are especially needed for underserved and racial and ethnic minority populations, who are disproportionately affected by the pandemic.^13-21^ In cases reported to the US Centers for Disease Control and Prevention, 33% of patients were Hispanic and 22% were Black, whereas these groups make up 18% and 13% of the US population, respectively.^14^ This is also a global issue; among 17 million patients in England, all non-White ethnic patients were found to have a higher risk of Covid-19–related death than White patients.^15^ Greater inclusion of minorities and underserved populations in clinical trials of Covid-19 therapies is needed; these populations are often not a focus of clinical trial recruitment and have been underrepresented in Covid-19 trials.^22^ Clinical data on these specific populations are urgently needed to improve patient outcomes.

Covid-19 may be associated with a dysregulated immune response and hyperinflammation, which can lead to acute respiratory distress syndrome and multiorgan failure.^4,23,24^ Studies have found that increased levels of interleukin-6 (IL-6) were associated with severe Covid-19 and increasing IL-6 levels correlated with increased likelihood of needing mechanical ventilation and mortality.^25-27^ The anti–IL-6 receptor monoclonal antibody tocilizumab, which is approved for multiple inflammatory diseases,^28,29^ has been found to improve outcomes in patients with Covid-19 pneumonia in observational studies in the US and globally.^30-34^ Although the randomized, placebo-controlled COVACTA trial—which enrolled patients with a range of disease severity, including 38% of patients receiving mechanical ventilation—did not meet its primary endpoint, its results suggested that tocilizumab may have a positive effect on recovery time and need for intensive care, which warrants further investigation.^35^

The global phase III EMPACTA (Evaluating Minority Patients with Actemra) clinical trial investigated the safety and efficacy of tocilizumab in hospitalized, nonventilated patients with Covid-19 pneumonia, with an emphasis on enrolling high-risk and racial and ethnic minority populations.

## PATIENTS AND METHODS

### Trial Design and Oversight

EMPACTA is a randomized, double-blind, placebo-controlled, phase III study to evaluate the safety and efficacy of tocilizumab in hospitalized, nonventilated patients with Covid-19 pneumonia (ClinicalTrials.gov NCT04372186). Inclusion of global study sites enrolling high-risk and minority populations was emphasized to enhance the understanding of the clinical profile of tocilizumab in these patients and allow access to underserved and minority populations, which are not commonly represented in clinical trials.

Patients ≥18 years of age (with no upper age limit) hospitalized with Covid-19 pneumonia confirmed by a positive polymerase chain reaction test and radiographic imaging were eligible. Patients had a blood oxygen saturation <94% on ambient air but were excluded if they required continuous positive airway pressure, bilevel positive airway pressure, or mechanical ventilation. Patients were receiving standard care per local practice, which could include antiviral treatment, limited systemic corticosteroids (≤1 mg/kg methylprednisolone or equivalent recommended) and supportive care. Patients were excluded if progression to death was imminent and inevitable within 24 hours as determined by the treating physician or they had active tuberculosis or suspected active bacterial, fungal, or viral infection (other than SARS-CoV-2 or well-controlled HIV). Patients with comorbidities were not excluded unless the investigator determined it would preclude safe patient participation.

All patients or their legally authorized representative gave informed consent. This study was conducted in accordance with the International Council for Harmonization E6 guideline for good clinical practice and the Declaration of Helsinki or local regulations, whichever afforded greater patient protection. Institutional review boards or ethics committees approved the protocol. The sponsor (Genentech, Inc.) designed the study, conducted it per the protocol, collected the data, and performed the analyses. All authors vouch for the accuracy and completeness of the data and for the trial’s fidelity to the protocol. All manuscript drafts were prepared by the authors, with editorial and writing assistance funded by the sponsor.

Patients were randomized (2:1) to intravenous tocilizumab (8 mg/kg, maximum 800 mg) or placebo, both plus standard care through permuted-block randomization and an interactive voice or web-based response system. Randomization was stratified by country (United States, Mexico, Kenya, South Africa, Peru, Brazil) and age (≤60 and >60 years). If a patient’s clinical signs or symptoms worsened or did not improve (reflected by sustained fever or worsening status on the 7-category ordinal scale), an additional infusion could be administered 8 to 24 hours after the first.

Efficacy was evaluated by Day 28, and patients were followed for a total of 60 days; patients discharged prior to Day 28 were considered study completers and followed weekly up to Day 28 with a safety follow-up by Day 60.

### Outcome Measures

The primary efficacy endpoint was cumulative proportion of patients requiring mechanical ventilation (mechanical invasive ventilation or extracorporeal membrane oxygen) or who had died by Day 28. Key secondary efficacy endpoints evaluated up to or by Day 28 were time to hospital discharge or ready for discharge based on a 7-category ordinal scale (Table S1 in the **Supplementary Appendix**); time to a ≥2 category improvement in clinical status relative to baseline on a 7-category ordinal scale (for patients in category 2 at baseline, having a clinical status of category 1 was considered meeting the threshold); time to clinical failure (time to death, mechanical ventilation, intensive care unit [ICU] admission [or 2-category worsening in the 7-category ordinal scale from baseline for patients already admitted into the ICU at study enrollment], or withdrawal [whichever occurred first]); and mortality rate. The primary efficacy endpoint was also analyzed by race/ethnicity subgroups.

Incidence and severity of adverse events, determined by the National Cancer Institute Common Terminology Criteria for Adverse Events v5.0, were evaluated.

### Statistical Analysis

A modified intent-to-treat (mITT, all randomized patients who received any study medication) population of 379 patients with 2:1 randomization was estimated to provide at least 80% power to detect a 15% difference between groups for the primary objective using a log-rank test assuming cumulative event rate (death or requiring mechanical ventilation) of 25% and 40% in the tocilizumab and placebo groups, respectively. Efficacy analyses were performed on the mITT population grouped according to randomization. Analyses were stratified by age group (≤60 or >60 years).

The primary endpoint was estimated using the Kaplan-Meier method, and cumulative incidence curves were compared between treatment groups using the stratified log-rank test. The stratified Cox proportional hazard model was used to estimate the hazard ratio (tocilizumab over placebo) and 95% confidence interval (CI) between treatment arms.

Primary and key secondary endpoints were evaluated in a hierarchical manner to control the overall study-wide type 1 error rate at the 5% significance level. If the primary endpoint reached statistical significance at the 2-sided 5% significance level, the key secondary endpoints were tested in the following predefined order: (1) time to hospital discharge/ready for discharge, (2) time to improvement in clinical status, (3) time to clinical failure endpoint, and (4) mortality. Time-to-event secondary endpoints were compared between treatment groups using the Kaplan-Meier approach. In the analyses for time to hospital discharge/ready for discharge and for time to improvement in clinical status, patients who had died or discontinued from study prior to hospital discharge/ready for discharge or prior to achieving improvement in clinical status were censored by Day 28 or date of the last ordinal scale assessment, respectively. In the analysis of the time to clinical failure endpoint, patients not experiencing clinical failure on or prior to Day 28 were censored at the last contact date or Day 28, whichever was earlier. Mortality by Day 28 was compared between treatment arms using the Cochran-Mantel-Haenszel test adjusted for age. For subgroup analyses, see the **Supplementary Appendix**.

Safety was assessed in the safety evaluable population, which was all patients who received any study medication; patients were grouped by the actual treatment received.

## RESULTS

### Patients

Overall, 389 patients from 6 countries were randomized, with 1 patient randomized in error, and 377 received study treatment (**Figure 1**). In the mITT population, 249 patients received tocilizumab plus standard of care and 128 received placebo plus standard of care. The safety population included 250 and 127 patients, respectively, because 1 patient randomized to placebo received tocilizumab and 11 patients did not receive study treatment. Study drug exposure is provided in Table S2 in the **Supplementary Appendix**. Overall, 225 patients (90.4%) completed the study in the tocilizumab arm and 115 (89.8%) in the placebo arm; excluding those who died, no tocilizumab patients and 2 placebo patients (1.6%) discontinued before Day 28. Median follow-up time was 10 days in both treatment arms.

**Figure 1.**
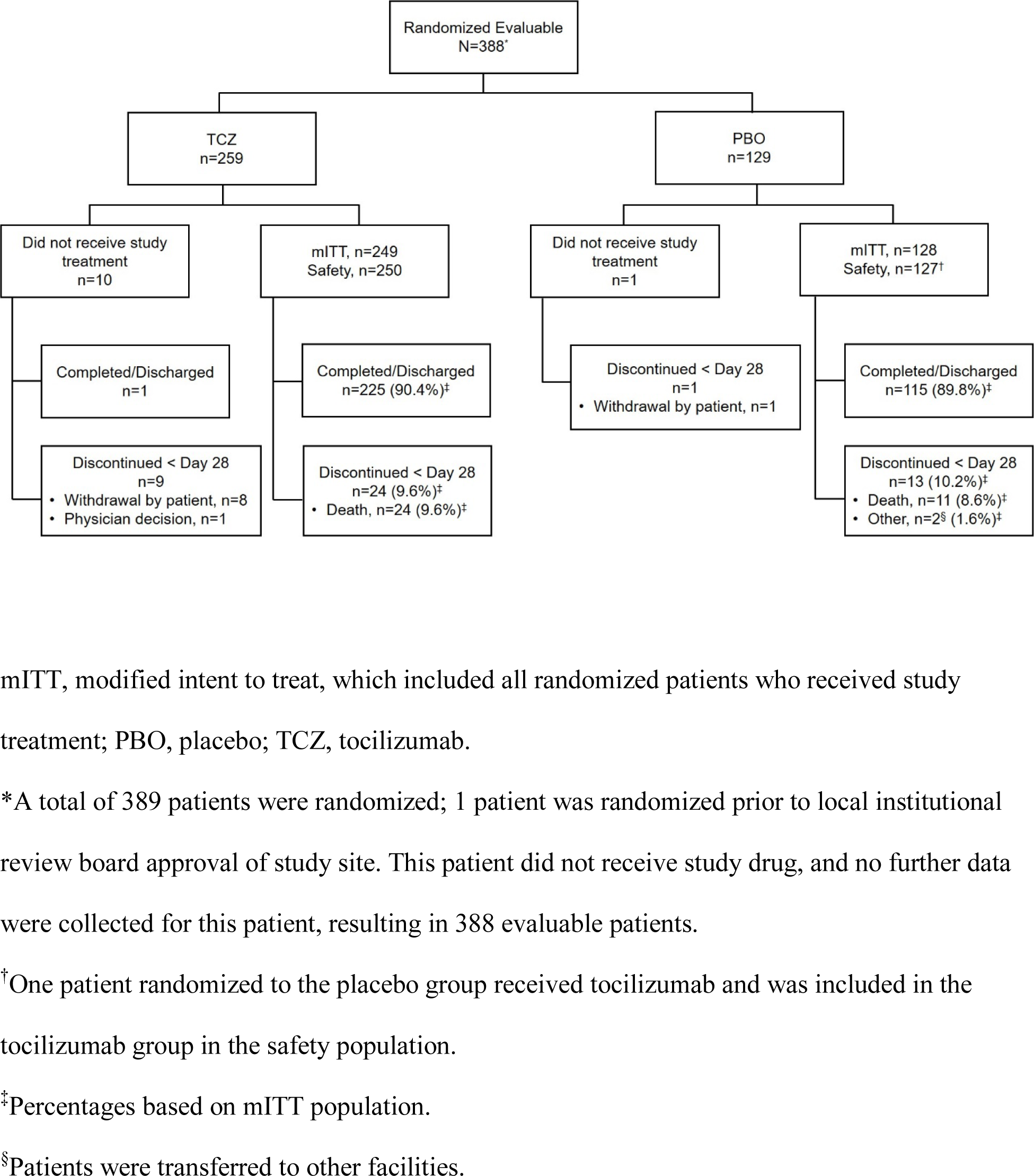
Patient Disposition.

Baseline demographics and disease characteristics were generally balanced between treatment arms (**Table 1**). In the tocilizumab and placebo arms, 60.2% and 57.0% of patients were male, respectively, and mean (± SD) age was 56.0 ±14.3 years and 55.6 ±14.9 years, respectively. In the tocilizumab and placebo arms, 143 patients (57.4%) and 68 (53.1%) were Hispanic/Latino, respectively, 35 patients (14.1%) and 21 (16.4%) were Black/African American, respectively, and 33 (13.3%) and 15 (11.7%) were American Indian/Alaska Native, respectively. In the tocilizumab and placebo arms, use of systemic corticosteroids (200 patients [80.3%] and 112 [87.5%]) and antiviral treatment (196 [78.7%] and 102 [79.7%]) in the 7 days prior to or during the study), respectively, were similar. In the tocilizumab and placebo arms, 55.4% and 67.2% of patients received dexamethasone, respectively, and 52.6% and 58.6% received remdesivir, respectively.

**Table 1.** Baseline Patient Demographics and Disease Characteristics (modified intent-to-treat population)*

|  | **Tocilizumab**<br>**n=249** | **Placebo**<br>**n=128** | **All Patients**<br>**N=377** |
| --- | --- | --- | --- |
| Sex, n (%) |  |  |  |
| Male | 150 (60.2) | 73 (57.0) | 223 (59.2) |
| Female | 99 (39.8) | 55 (43.0) | 154 (40.8) |
| Age, mean (SD), years | 56.0 (14.3) | 55.6 (14.9) | 55.9 (14.4) |
| Age, median (range), years | 57.0 (24-95) | 56.0 (20-89) | 57.0 (20-95) |
| Age group, n (%), years |  |  |  |
| 18-64 | 178 (71.5) | 93 (72.7) | 271 (71.9) |
| 65-84 | 70 (28.1) | 33 (25.8) | 103 (27.3) |
| ≥ 85 | 1 (0.4) | 2 (1.6) | 3 (0.8) |
| Weight, mean (SD), kg | 89.57 (23.73) | 94.44 (25.95) | 91.22 (24.58) |
| Weight, median (range), kg | 85.00<br>(45.4-171.8) | 90.05<br>(44.0-201.0) | 86.00<br>(44.0-201.0) |
| BMI |  |  |  |
| n | 240 | 122 | 362 |
| BMI, mean (SD) | 32.03 (7.86) | 33.05 (7.18) | 32.38 (7.64) |
| BMI, median (range) | 30.60<br>(18.4-66.2) | 31.95<br>(17.2-55.7) | 30.90<br>(17.2-66.2) |
| Race/ethnicity, n (%)<sup>†</sup> |  |  |  |
| Hispanic or Latino | 143 (57.4) | 68 (53.1) | 211 (56.0) |
| American Indian or Alaska Native | 33 (13.3) | 15 (11.7) | 48 (12.7) |
| Black or African American | 35 (14.1) | 21 (16.4) | 56 (14.9) |
| Non-Hispanic White | 28 (11.2) | 20 (15.6) | 48 (12.7) |
| Unknown/other | 10 (4.0) | 4 (3.1) | 14 (3.7) |
| Country group, n (%) |  |  |  |
| US | 201 (80.7) | 103 (80.5) | 304 (80.6) |
| Ex-US<sup>‡</sup> | 48 (19.3) | 25 (19.5) | 73 (19.4) |
| Smoking history, n (%) |  |  |  |
| Never | 192 (77.1) | 99 (77.3) | 291 (77.2) |
| Current | 16 (6.4) | 6 (4.7) | 22 (5.8) |
| Former | 41 (16.5) | 23 (18.0) | 64 (17.0) |
| Ordinal scale for clinical status, n (%)<sup>§</sup> |  |  |  |
| 2 | 24 (9.6) | 11 (8.6) | 35 (9.3) |
| 3 | 161 (64.7) | 81 (63.3) | 242 (64.2) |
| 4 | 64 (25.7) | 36 (28.1) | 100 (26.5) |
| Elevated CRP, n (%) |  |  |  |
| n | 232 | 120 | 352 |
| Yes | 190 (81.9) | 104 (86.7) | 294 (83.5) |
| No | 42 (18.1) | 16 (13.3) | 58 (16.5) |
| CRP, mg/L |  |  |  |
| n | 191 | 99 | 290 |
| Mean (SD) | 151.94 (177.22) | 202.80 (404.92) | 169.30 (277.18) |
| Median (range) | 123.40<br>(2.5-2099.0) | 143.40<br>(9.0-3776.0) | 136.05<br>(2.5-3776.0) |
| hs-CRP, mg/L |  |  |  |
| n | 41 | 21 | 62 |
| Mean (SD) | 98.23 (111.36) | 88.46 (75.01) | 94.92 (100.00) |
| Median (range) | 68.25<br>(0.1-494.7) | 76.40<br>(2.0-290.7) | 70.85<br>(0.1-494.7) |
| D-dimer, ug/mL FEU |  |  |  |
| n | 223 | 116 | 339 |
| Mean (SD) | 423.54<br>(1095.14) | 472.69<br>(1392.65) | 440.36<br>(1203.39) |
| Median (range) | 1.60<br>(0.2-8873.0) | 1.21<br>(0.2-10384.0) | 1.55<br>(0.2-10384.0) |
| Ferritin levels, pmol/L |  |  |  |
| n | 214 | 111 | 325 |
| Mean (SD) | 2305.96<br>(3431.54) | 3585.47<br>(12533.99) | 2742.96<br>(7838.84) |
| Median (range) | 1408.19<br>(29.2-38482.1) | 1353.14<br>(110.1-122328.9) | 1397.18<br>(29.2-122328.9) |
| Days from first Covid-19 symptom at baseline |  |  |  |
| n | 248 | 127 | 375 |
| Mean (SD) | 7.82 (4.36) | 8.55 (6.07) | 8.07 (5.01) |
| Median (range) | 8.00 (0.0-31.0) | 8.00 (0.0-36.0) | 8.00 (0.0-36.0) |
| Days from Covid-19 diagnosis |  |  |  |
| Mean (SD) | 2.25 (2.44) | 2.13 (2.24) | 2.21 (2.37) |
| Median (range) | 1.00 (0.0-14.0) | 2.00 (0.0-12.0) | 2.00 (0.0-14.0) |
| ICU admission status at baseline, n (%) |  |  |  |
| Yes | 36 (14.5) | 22 (17.2) | 58 (15.4) |
| No | 213 (85.5) | 106 (82.8) | 319 (84.6) |
| Comorbidities, n (%)<sup>¶ #</sup> |  |  |  |
| n | 250 | 127 | 377 |
| Patients with ≥1 comorbidity | 191 (76.4) | 96 (75.6) | 287 (76.1) |
| Asthma | 27 (10.8) | 16 (12.6) | 43 (11.4) |
| Atrial fibrillation | 6 (2.4) | 6 (4.7) | 12 (3.2) |
| Chronic obstructive pulmonary disease | 12 (4.8) | 5 (3.9) | 17 (4.5) |
| Diabetes | 105 (42.0) | 48 (37.8) | 153 (40.6) |
| Hyperlipidemia | 70 (28.0) | 34 (26.8) | 104 (27.6) |
| Hypertension | 119 (47.6) | 63 (49.6) | 182 (48.3) |
| Myocardial infarction | 4 (1.6) | 3 (2.4) | 7 (1.9) |
| Obesity | 54 (21.6) | 38 (29.9) | 92 (24.4) |
| Stroke | 10 (4.0) | 3 (2.4) | 13 (3.4) |

BMI, body mass index; CRP, C-reactive protein; FEU, fibrinogen equivalent units; hs, high sensitivity; ICU, intensive care unit.
\* Unless otherwise indicated.
<sup>†</sup>Self-reported.
<sup>‡</sup>Mexico, Kenya, South Africa, Peru, Brazil.
<sup>§</sup>7-category ordinal scale: 1, discharged (or ready for discharge as evidenced by normal body temperature and respiratory rate, and stable oxygen saturation on ambient air or $\leq 2$ L supplemental oxygen); 2, non-ICU hospital ward (or ready for hospital ward), not requiring supplemental oxygen; 3, non-ICU hospital ward (or ready for hospital ward), requiring supplemental oxygen; 4, ICU or non-ICU hospital ward, requiring noninvasive ventilation or
high-flow oxygen; 5, ICU, requiring intubation and mechanical ventilation; 6, ICU, requiring extracorporeal membrane oxygenation or mechanical ventilation and additional organ support; 7, death.
‖ Patients with CRP >50 mg/L or hs-CRP >3 mg/L at baseline.
¶ Safety population.

### Primary Efficacy Outcome

The primary efficacy comparison was statistically significant; the cumulative proportion of patients requiring mechanical ventilation or who had died by Day 28 was 12.0% (95% CI, 8.52% to 16.86%) with tocilizumab and 19.3 % (95% CI, 13.34% to 27.36%) with placebo (hazard ratio, 0.56 [95% CI, 0.33 to 0.97]); log-rank P=0.0360; **Table 2, Figure 2**).

**Table 2.** Primary and Key Secondary Efficacy Endpoints (modified intent-to-treat population)

|  | **Tocilizumab**<br>**n=249** | **Placebo**<br>**n=128** | **Comparison** |  |  |
| --- | --- | --- | --- | --- | --- |
|  |  |  | **Hazard**<br>**Ratio**<br>**(95% CI)** | **Weighted**<br>**Difference,**<br>**% (95% CI)** | **P value** |
| Cumulative proportion (95% CI)* of patients requiring mechanical ventilation or who had died by Day 28 | 12.0<br>(8.52-16.86) | 19.3<br>(13.34-27.36) | 0.56<br>(0.33-0.97) | — | 0.0360<sup>†</sup> |
| Time to hospital discharge or ready for discharge, median (95% CI),<sup>‡</sup> days | 6.0<br>(6.0-7.0) | 7.5<br>(7.0-9.0) | 1.16<br>(0.91-1.48) | — | § |
| Time to improvement in ordinal clinical status to Day 28, mITT population, median (95% CI), <sup>‡</sup> days | 6.0<br>(6.0-7.0) | 7.0<br>(6.0-9.0) | 1.15<br>(0.90-1.48) | — | § |
| Time to clinical failure, median (95% CI),<sup>‡</sup> days | NE | NE | 0.55<br>(0.33-0.93) | — | § |
| Mortality by Day 28, n (%) [95% CI]<sup>¶#</sup> | 26 (10.4)<br>[7.2-14.9] | 11 (8.6)<br>[4.9-14.7] | — | 2.0<br>(-5.2-7.8) | § |

ICU, intensive care unit; mITT, modified intent to treat; NE, not estimable.
\*Cumulative proportion of patients was estimated using the Kaplan-Meier method and compared between treatment groups using the stratified log-rank test with age group ( $\leq 60$ , $>60$ years) as a stratification factor. The stratified Cox proportional hazard model with age group as a stratification factor ( $\leq 60$ , $>60$ years) was used to estimate the hazard ratio and 95% CI between treatment arms.
<sup>†</sup>P values were calculated with the log-rank test.
<sup>‡</sup>Time-to-event secondary endpoints were estimated using the Kaplan-Meier approach.
<sup>§</sup>Significance testing was performed hierarchically to control for study-wide type I error rate at a 5% significance level. Time-to-event secondary endpoints were compared between treatment groups using the Kaplan-Meier approach. P value not presented because first secondary endpoint failed to reach significance.
<sup>¶</sup>7-category ordinal scale: 1, discharged (or ready for discharge as evidenced by normal body temperature and respiratory rate, and stable oxygen saturation on ambient air or $\leq 2$ L supplemental oxygen); 2, non-ICU hospital ward (or ready for hospital ward), not requiring supplemental oxygen; 3, non-ICU hospital ward (or ready for hospital ward), requiring
supplemental oxygen; 4, ICU or non-ICU hospital ward, requiring noninvasive ventilation or high-flow oxygen; 5, ICU, requiring intubation and mechanical ventilation; 6, ICU, requiring extracorporeal membrane oxygenation or mechanical ventilation and additional organ support; 7, death.
<sup>¶</sup>The Wilson method was used to estimate the 95% CI for the observed proportion. The Cochran-Mantel-Haenszel weighting approach with age group as the stratification factor ( $\leq 60$ , $>60$ years) was used to calculate the weighted difference in percentages. The Newcombe method was used to estimate the 95% CI for the weighted difference.
<sup>#</sup>Included all deaths reported from ordinal scale scoring, adverse events reporting and public death records, during hospital stay and after hospital discharge, through Day 28.

**Figure 2.**
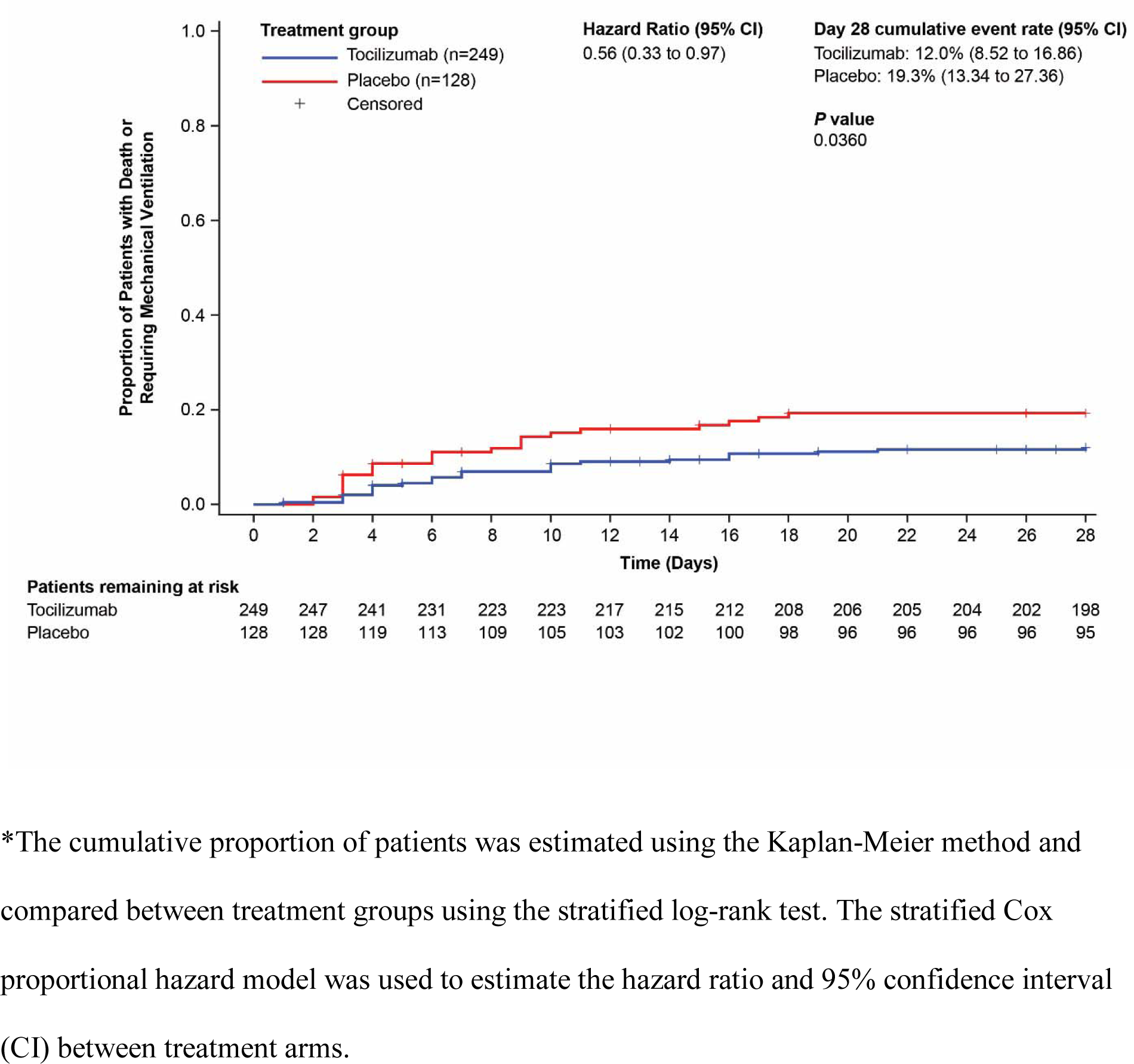
Cumulative Proportion Plot of Time to Requiring Mechanical Ventilation or Death by Day 28 (modified-intent-to-treat population)*

### Secondary Outcomes

Median (95% CI) time to hospital discharge/ready for discharge up to Day 28 was 6.0 days (6.0 to 7.0) with tocilizumab and 7.5 days (7.0 to 9.0) with placebo (hazard ratio, 1.16 [95% CI, 0.91 to 1.48]); **Table 2, Figure S1**). Median (95% CI) time to improvement in ordinal clinical status up to Day 28 was 6.0 days (6.0 to 7.0) with tocilizumab and 7.0 days (6.0 to 9.0) with placebo (hazard ratio, 1.15 [95% CI, 0.90 to 1.48]; **Table 2, Figure S2**). Median (95% CI) time to clinical failure up to Day 28 was not reached in either group (hazard ratio, 0.55 [95% CI, 0.33 to 0.93]; **Table 2, Figure S3**). Mortality (95% CI) by Day 28 was 10.4% (7.2% to 14.9%) in the tocilizumab arm and 8.6% (4.9% to 14.7%) in the placebo arm (weighted difference, 2% [95% CI, –5.2% to 7.8%)]; **Table 2**).

### Exploratory Outcomes

Primary efficacy analysis results by race/ethnicity were consistent with the result for all patients (**Figure S4**).

### Safety

Overall, adverse events were reported in 50.8% of 250 patients and 52.8% of 127 patients in the tocilizumab and placebo arms, respectively, through Day 60 (the data cutoff was September 30, 2020) and serious adverse events in 15.2% and 19.7% (**Table 3**). Deaths occurred in 29 patients (11.6%) in the tocilizumab arm and 15 (11.8%) in the placebo arm. Through Day 60, 13 patients receiving tocilizumab (5.2%) had 16 serious infections and 9 patients receiving placebo (7.1%) had 11.

**Table 3.** Safety Through Day 60 (safety population)

|  | **Tocilizumab**<br>**n=250** | **Placebo**<br>**n=127** | **All Patients**<br>**N=377** |
| --- | --- | --- | --- |
| Total adverse events, n | 357 | 187 | 544 |
| Patients with $\geq 1$ adverse event, n (%) | 127 (50.8) | 67 (52.8) | 194 (51.5) |
| Total deaths, n (%) | 29 (11.6) | 15 (11.8) | 44 (11.7) |
| Total number of patients discontinued from study due to an adverse event, n (%) | 0 | 0 | 0 |
| Patients with $\geq 1$ event, n (%) | | | |
| Adverse event with fatal outcome | 28 (11.2) | 13 (10.2) | 41 (10.9) |
| Serious adverse event | 38 (15.2) | 25 (19.7) | 63 (16.7) |
| Serious adverse event leading to withdrawal from study (excluding serious adverse events leading to death) | 0 | 0 | 0 |
| Serious adverse event leading to dose modification or interruption | 0 | 0 | 0 |
| Treatment-related* serious adverse event | 3 (1.2) | 0 | 3 (0.8) |
| Adverse event leading to withdrawal from study (excluding adverse events leading to death) | 0 | 0 | 0 |
| Adverse event leading to dose modification or interruption | 1 (0.4) | 0 | 1 (0.3) |
| Treatment-related* adverse event | 32 (12.8) | 5 (3.9) | 37 (9.8) |
| Treatment-related* adverse event leading to withdrawal from treatment | 0 | 0 | 0 |
| Treatment-related* adverse event leading to dose modification or interruption | 0 | 0 | 0 |
| Grades 3 to 5 adverse event (at greatest intensity)<sup>†</sup> | 46 (18.4) | 31 (24.4) | 77 (20.4) |
| Infection adverse event | 25 (10.0) | 16 (12.6) | 41 (10.9) |
| Serious infection adverse event | 13 (5.2) | 9 (7.1) | 22 (5.8) |
| Total number of events | 16 | 11 | 27 |
| Events with incidence of >1%<br>in either treatment arm |  |  |  |
| Septic shock | 5 (2.0) | 3 (2.4) | 8 (2.1) |
| Covid-19 pneumonia | 2 (0.8) | 3 (2.4) | 5 (1.3) |
| Pneumonia | 0 | 3 (2.4) | 3 (0.8) |
| Pneumonia bacterial | 0 | 2 (1.6) | 2 (0.5) |

\*As determined by the investigator.
† Incidence and severity of adverse events as determined by the National Cancer Institute Common Terminology Criteria for Adverse Events v5.0, were evaluated.

## DISCUSSION

The phase III EMPACTA trial of tocilizumab in hospitalized nonventilated patients with Covid-19 pneumonia emphasized enrolling high-risk and racial and ethnic minority patients, who are disproportionately affected by Covid-19 and often underrepresented in clinical trials.^13-22^ Overall, >25% of patients were over age 65, >75% had ≥1 comorbidity and >80% were a racial or ethnic minority. This trial demonstrated significant reduction in the likelihood of progressing to mechanical ventilation or death by Day 28 in patients receiving tocilizumab plus standard care vs placebo plus standard care. When all-cause mortality alone was evaluated as a secondary endpoint, no mortality benefit was observed; however, this study was not specifically designed or powered to detect differences in mortality, making it difficult to draw conclusions based on these data. Time to clinical failure up to Day 28 favored tocilizumab over placebo.

Healthcare disparities in Covid-19 are a critical issue as studies continue to show that racial and ethnic minority populations are disproportionately affected by the pandemic.^13-21^ The role of race and ethnicity in Covid-19 is complex and not fully understood; social determinants of health, socioeconomic factors, and historic and structural inequities may play a role but do not completely explain the observations in the body of evidence, and further research is needed.^16,18,21,36^ EMPACTA directly addressed the discrepancy between the overrepresentation of racial and ethnic minorities with Covid-19 disease and their underrepresentation in Covid-19 trials^22^ by prioritizing sites that provide care to underserved and minority populations while continuing to enroll all eligible patients. As a result, 84% were either Hispanic/Latino, Black/African American, or American Indian/Alaska Native. Although the analysis was exploratory, the primary efficacy outcomes based on race were consistent with the outcome in the overall patient population.

Preventing progression to mechanical ventilation, which may greatly alter patient outcomes and healthcare resource availability, is critical for potential Covid-19 therapies. Mechanical ventilation is administered as disease severity increases and patients experience respiratory distress and failure. In an observational study of critically ill patients with Covid-19, 76% received mechanical ventilation and the mortality rate among these patients was 35.7%.^8^ In a global literature survey of hospitalized patients with Covid-19, 63% of those in the ICU received mechanical ventilation and the mortality rate for these patients was 59%.^9^ In addition to clinical decision-making, one of the main factors determining whether patients receive mechanical ventilation is availability of resources. Critical shortages of ventilators have occurred globally during the pandemic, underscoring the need for effective therapies that preserve this limited resource^37^; this is especially a concern in African countries.^38^ Reducing the proportion of patients who progress to requiring mechanical ventilation, as this study demonstrated in patients receiving tocilizumab, could help reduce the burden on critical care services, reduce direct healthcare costs, and ensure availability for the most critical patients.^39^

Besides EMPACTA, the efficacy of tocilizumab in Covid-19 has been examined in one other randomized placebo-controlled trial, COVACTA.^35^ COVACTA included patients across a spectrum of disease severity, from baseline moderate hypoxia to invasive mechanical ventilation, while EMPACTA enrolled patients who were not ventilated at baseline and who were at an earlier stage of disease. Furthermore, in contrast with COVACTA, in EMPACTA most patients received concomitant corticosteroids or antivirals, which have become the mainstay of Covid-19 standard care,^40^ although these treatments were not used uniformly and patients may not have received a full treatment course. Corticosteroid and antiviral use was generally balanced between arms, and EMPACTA demonstrated the added clinical benefit of tocilizumab. In COVACTA, the median time to hospital discharge was 20 and 28 days in the tocilizumab and placebo arms, respectively, whereas in EMPACTA, it was 6 and 7.5 days, respectively.^35^ This may reflect patients who were more severely ill or different comorbid conditions at study baseline in COVACTA, as well as improvements in the standards of care and different proportions of patients receiving concomitant steroids and antivirals. Tocilizumab was associated with a reduced risk of invasive mechanical ventilation or death (adjusted hazard ratio, 0.61 [95% CI, 0.40–0.92]; P=0.020) in a retrospective study.^32^

EMPACTA demonstrated the efficacy and safety of tocilizumab plus standard care over placebo plus standard care in reducing the likelihood of progression to requiring mechanical ventilation or death among hospitalized nonventilated patients with Covid-19 pneumonia; this is the first placebo-controlled trial to demonstrate a likelihood of reduction in progression to mechanical ventilation in Covid-19. EMPACTA is also the first trial of its kind to emphasize diversity and ensure underserved populations are represented in clinical research and given the opportunity to receive novel therapies.

## Data Availability

Qualified researchers may request access to individual patient level data through the clinical study data request platform (https://vivli.org/). Further details on Roche's criteria for eligible studies are available here (https://vivli.org/members/ourmembers/). For further details on Roche's Global Policy on the Sharing of Clinical Information and how to request access to related clinical study documents, see here (https://www.roche.com/research_and_development/who_we_are_how_we_work/clinical_trials/our_commitment_to_data_sharing.htm).

https://vivli.org/members/ourmembers/

https://www.roche.com/research_and_development/who_we_are_how_we_work/clinical_trials/our_commitment_to_data_sharing.htm

## Acknowledgments

We thank the patients who participated in this trial, the clinical site investigators, and Claire Stedden, Ph.D., and Nicola Gillespie, DVM, of Health Interactions for medical writing assistance with an earlier version of the manuscript. This study was funded by Genentech, Inc.

