## Supplementary Appendix for "Tocilizumab in nonventilated patients hospitalized with Covid-19 pneumonia"

**Contents**

| Study investigators………………………………………………………….……………... | 2 |
| --- | --- |
| Methods for subgroup analyses ………………………………………….………………... | 4 |
| **Table S1.** 7-category ordinal scale …………………………………………..………...….. | 5 |
| **Table S2.** Study drug exposure in safety population…………………………………..….. | 6 |
| **Figure S1.** Time to hospital discharge/ready for discharge up to Day 28 ………………... | 7 |
| **Figure S2.** Time to improvement in ordinal clinical status up to Day 28………………… | 8 |
| **Figure S3.** Time to clinical failure up to Day 28……………………………………...…... | 9 |
| **Figure S4.** Subgroup analysis by race/ethnicity of the cumulative proportion of patients requiring mechanical ventilation or who died by Day 28…….…………………………… | 10 |

**Study investigators**

| **Study Site** | **Principal Investigator** |
| --- | --- |
| **United States** | |
| **Highland Hospital, Oakland, CA + San Leandro Hospital, San Leandro, CA** | Rachel Baden, MD |
| **Banner University Medical Center, Phoenix, AZ** | Marilyn Glassberg, MD |
| **Canton Potsdam Hospital, Potsdam, NY** | Eyal Kedar, MD |
| **Cape Fear Valley Medical Center, Fayetteville, NC** | Judith Borger, DO |
| **El Centro Regional Medical Center, El Centro, CA** | Jorge Robles, MD |
| **Elmhurst Hospital, Elmhurst, NY** | Emma Kaplan-Lewis, MD |
| **Flushing Hospital, Queens, NY** | Javeria Shakil, MD |
| **Harlem Hospital, New York, NY** | Sharon Mannheimer, MD |
| **Henry Ford Health System, Detroit, MI** | Mayur Ramesh, MD |
| **Holy Cross Hospital + Holy Cross Germantown Hospital, Bethesda, MD** | Miriam Cameron, MD |
| **Houston VA Medical Center, Houston, TX** | Barbara Trautner, MD |
| **Jamaica Hospital, New York, NY** | Khalid Gafoor, DO |
| **McAllen Medical Center, McAllen, TX** | Carlos Palacio Lascano, MD |
| **Miami VA Medical Center, Miami, FL** | Paola Lichtenberger, MD |
| **Novant Health Presbytarian Medical Center, Charlotte, NC + Novant Health Forsyth Medical Center, Winston-Salem, NC + Novant Health Rowan Medical Center, Salisbury, NC** | Alan Skarbnik, MD |
| **Ochsner, New Orleans, LA** | Julia Garcia-Diaz, MD |
| **San Juan Oncology Associates, Farmington, NM** | Jeffrey Neidhart, MD |
| **Sentara Norfolk General Hospital, Norfolk, VA** | Anthony Quaranta, MD |
| **Sharp Chula Vista Medical Center, Chula Vista, CA** | Michael Waters, MD |
| **St Barnabas Hospital, Bronx, NY** | Victoria Bengualid, MD |
| **St Joseph Hospital, Orange County, CA** | Timothy Byun, MD |
| **St Luke’s Boise Medical Center, Boise, ID** | Karen Miller, MD |
| **Temple University, Philadelphia, PA** | Gerard Criner, MD |
| **University of Arizona, Tucson, AZ** | Sachin Chaudhary, MD |
| **University of Miami Pulmonary, Miami, FL** | David De La Zerda, MD |
| **Valley Baptist Medical Center, Harlingen, TX** | Christopher Romero, MD |
| **Westchester General Hospital + Larkin Community Hospital, Miami, FL** | Luis Mendez-Mulet, MD |
| **Peru** | |
| **Hospital Maria Auxiliadora, Lima** | Maria Angelica Paredes Moreno, MD |
| **Hospital Militar Central, Lima** | Victoria Chavez Miñano, MD |
| **Hospital Nacional Cayetano Heredia, Lima** | Fernando Mejía Cordero, MD |
| **Hospital Nacional Hipolito Unanue, Lima** | Johan Azañero, MD |
| **Hospital Nacional Sergio E. Bernales, Lima** | Epifanio Sanchez Garavito, MD |
| **Brazil** | |
| **Centro Multidisciplinar de Estudos Clinicos CEMEC FMABC, São Paulo** | Adilson Joaquim Westheimer Cavalcante, MD |
| **Hospital E Maternidade Celso Pierro PUCCAMP, São Paulo** | Maria Lima, MD |
| **Hospital Municipal de Barueri Dr. Francisco Moran, São Paulo** | Ferdinando Lima Menezes, MD |
| **Kenya** | |
| **Aga Khan University Hospital, Nairobi** | Reena Shah, MScID, FRCP |
| **South Africa** | |
| **George Provincial Hospital, George** | Martha Mekebeb-Reuter, MD |
| **Mexico** | |
| **Hospital General de Culiacán, Culiacán** | Jorge Zamudio, MD |
| **Instituto Nacional de Ciencias Medicas y Nutrición, Mexico City** | Maria F. González-Lara, MD, MSc |

**Methods for subgroup analyses**

For the analysis by race/ethnicity subgroup, the unstratified Cox proportional hazard model was used to estimate the hazard ratio and 95% confidence interval (CI) between treatment arms.

**Table S1.** 7-Category Ordinal Scale

| **Category** | **Clinical Status** |
| --- | --- |
| 1 | Discharged (or ready for discharge as evidenced by normal body temperature and respiratory rate, and stable oxygen saturation on ambient air or ≤2L supplemental oxygen) |
| 2 | Non–ICU hospital ward (or ready for hospital ward), not requiring supplemental oxygen |
| 3 | Non–ICU hospital ward (or ready for hospital ward), requiring supplemental oxygen |
| 4 | ICU or non–ICU hospital ward, requiring noninvasive ventilation or high-flow oxygen |
| 5 | ICU, Requiring intubation and mechanical ventilation |
| 6 | ICU, requiring extracorporeal membrane oxygenation or mechanical ventilation and additional organ support (e.g., vasopressors or renal replacement therapy) |
| 7 | Death |

ICU, intensive care unit.

**Table S2.** Study Drug Exposure in Safety Population

|  | **Tocilizumab n=250** | **Placebo n=127** | **All Patients N=377** |
| --- | --- | --- | --- |
| Number of doses administered, n (%) |  |  |  |
| 1 | 182 (72.8) | 92 (72.4) | 274 (72.7) |
| 2 | 68 (27.2) | 35 (27.6) | 103 (27.3) |
| Time from first dose to second dose, hours* |  |  |  |
| n | 51 | 23 | 74 |
| Mean (SD) | 19.26 (3.40) | 20.78 (6.20) | 19.73 (4.47) |
| Median (range) | 20.15 (10.2-26.2) | 20.15 (10.3-43.9) | 20.15 (10.2-43.9) |

*Time from first dose to second dose was calculated as the start time of the second dose minus the end time of the first dose. This measure was not calculated for patients with missing time information in dosing records.

**Figure S1.** Kaplan-Meier plot of time to hospital discharge/ready for discharge up to Day 28* (modified-intent-to-treat population)

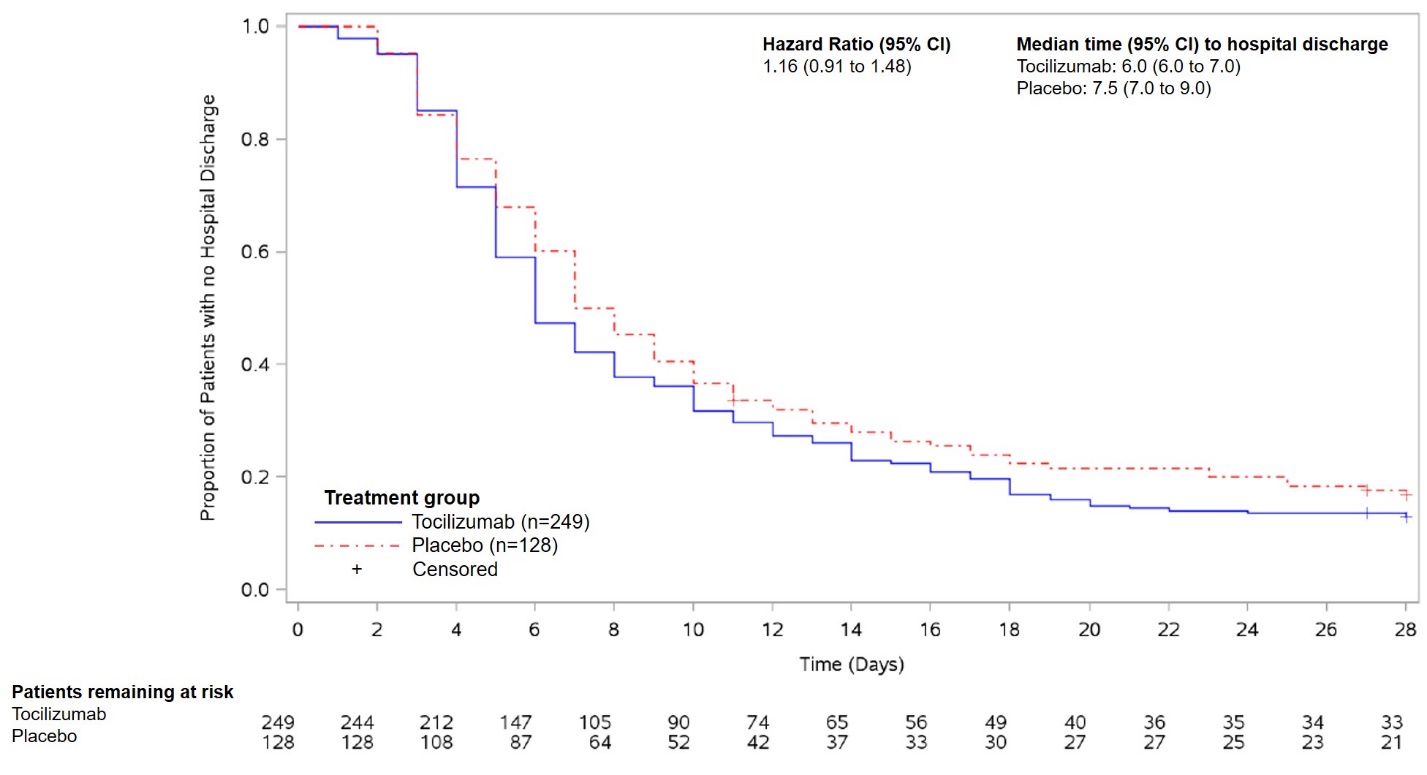

Significance testing was performed hierarchically to control for study-wide type I error rate at a 5% significance level. P value not presented because first secondary endpoint failed to reach significance.

The stratified Cox proportional hazard model was used to estimate the hazard ratio and 95% confidence interval (CI) between treatment arms.

*Patients who died or discontinued from study prior to hospital discharge or ready for discharge were censored at Day 28 or date of the last ordinal scale assessment.

**Figure S2.** Kaplan-Meier plot of time to improvement in ordinal clinical status^*^ up to Day 28^†^ (modified-intent-to-treat population)

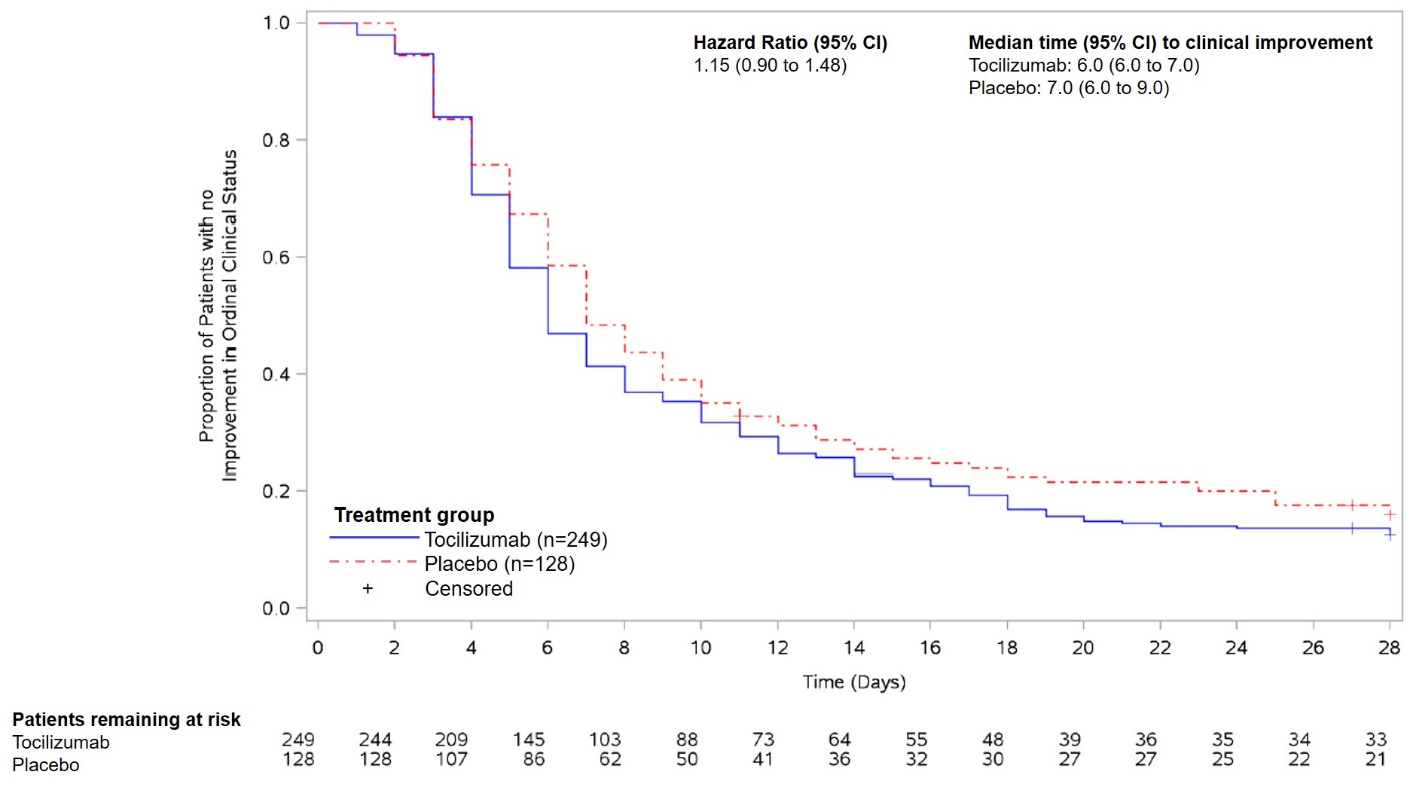

Significance testing was performed hierarchically to control for study-wide type I error rate at a 5% significance level. P value not presented because first secondary endpoint failed to reach significance.

The stratified Cox proportional hazard model was used to estimate the hazard ratio and 95% confidence interval (CI) between treatment arms.

*7-category ordinal scale: 1, discharged (or ready for discharge as evidenced by normal body temperature and respiratory rate, and stable oxygen saturation on ambient air or ≤2L supplemental oxygen); 2, non–intensive care unit (ICU) hospital ward (or ready for hospital ward), not requiring supplemental oxygen; 3, non–ICU hospital ward (or ready for hospital ward), requiring supplemental oxygen; 4, ICU or non–ICU hospital ward, requiring noninvasive ventilation or high-flow oxygen; 5, ICU, requiring intubation and mechanical ventilation; 6, ICU, requiring extracorporeal membrane oxygenation or mechanical ventilation and additional organ support; 7, death.

^†^Patients who died or discontinued from study prior to achieving improvement in clinical status were censored at Day 28 or date of the last ordinal scale assessment.

**Figure S3.** Cumulative proportion plot of time to clinical failure up to Day 28* (modified-intent-to-treat population)

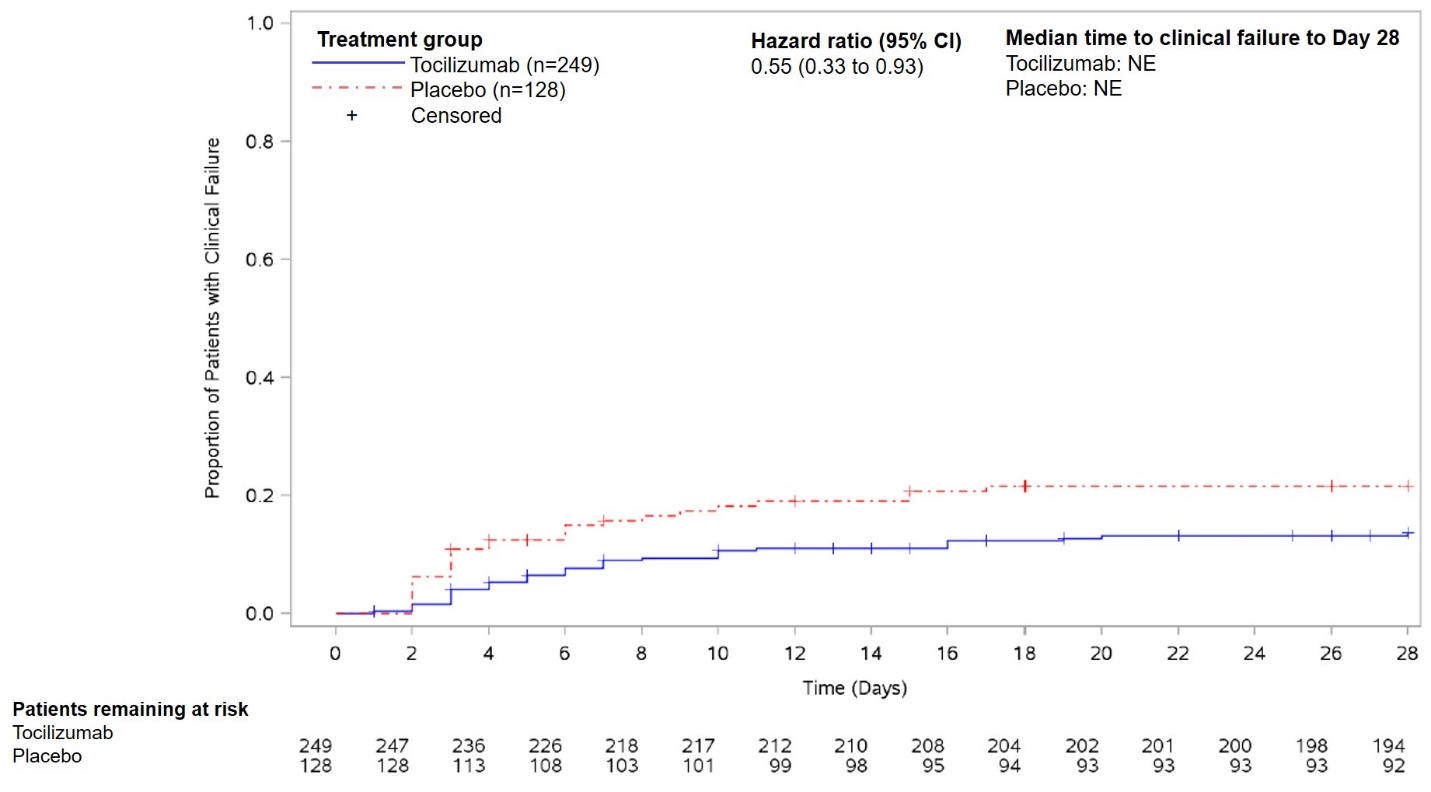

NE, not estimable.

Significance testing was performed hierarchically to control for study-wide type I error rate at a 5% significance level. P value not presented because first secondary endpoint failed to reach significance.

The stratified Cox proportional hazard model was used to estimate the hazard ratio and 95% confidence interval (CI) between treatment arms.

*Patients not experiencing any clinical failure on or prior to Day 28 were censored at the last contact date or Day 28, whichever was earlier.

**Figure S4.** Forest plot of hazard ratio for time to death or requiring mechanical ventilation up to Day 28 by race/ethnicity subgroup (modified-intent-to-treat population)

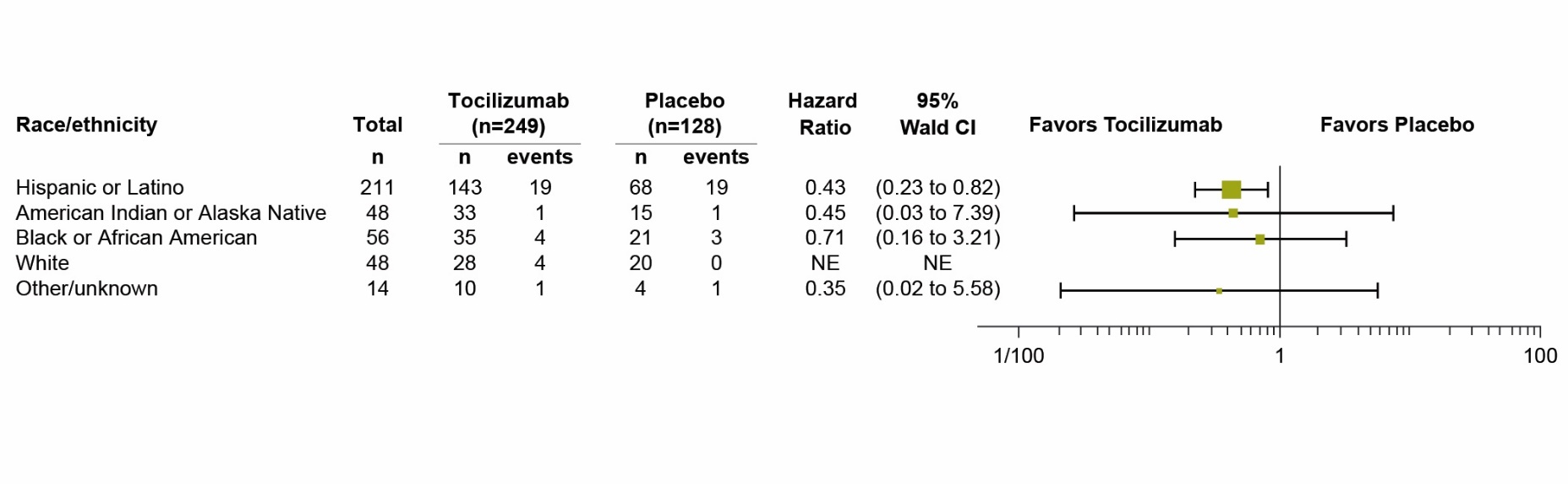

The unstratified Cox proportional hazard model was used to estimate the hazard ratio and 95% confidence interval (CI) between treatment arms. Unstratified hazard ratios are displayed. Hazard ratio < 1 favors tocilizumab.
